# Imaging Modalities for Left Atrial Appendage Occlusion: Post-Procedural Transesophageal Echocardiography vs Cardiac Computed Tomography

**DOI:** 10.1101/2025.07.14.25331485

**Authors:** Sujoy Khasnavis, Kusha Rahgozar, Michael Grushko, Jay Gross

**Author notes:** Corresponding address: Montefiore Medical Center, 111 E 210th St, Bronx, NY 10467 Address correspondence to: Sujoy Khasnavis, MD.

## Abstract

**Background:** The left atrial appendage (LAA) is considered a primary site for thrombus formation due to blood stagnation in the appendage. Left atrial appendage occlusion (LAAO) at the orifice is often a viable approach for preventing thromboembolism. Post LAAO evaluation with transesophageal echocardiography (TEE) is considered the standard modality to examine for potential complications. The use of cardiac computed tomography (CCTA) has recently emerged as a potential alternative to TEE.

**Methods:** Databases of Pubmed, Embase, Cochrane Library, and Web of Science were investigated for review studies on post LAAO evaluation with TEE and CCTA.

**Results:** TEE allows assessment of device position, thrombi, peri-device leaks (PDLs), biventricular function, and valve function with clinically informative images. CCTA also allows prediction of device behavior, position, and stability. CCTA further produces high-resolution images for evaluation of heart, vasculature, and devices. It more accurately detects device thrombi and PDLs, thereby better protecting against thromboembolic complications. CCTA involves fewer personnel and potentially lower costs. Among the disadvantages of TEE are that it is invasive, risks esophageal perforations and respiratory compromise, and requires many personnel which add to costs. For CCTA there is a risk of worsening renal function with resultant kidney injury which is of particular concern in chronic kidney disease (CKD). Moreover, CCTA utilizes x-ray radiation which increases risk of long term cancer.

**Conclusion:** While head to head comparisons are limited, CCTA has some key advantages over TEE in post LAAO evaluation such as higher rates of detection for PDLs, LAATs, and device positioning as well as ease and speed of performance. Further investigations will help determine the patient level outcomes, population outcomes, and the cost benefits of CCTA vs TEE.

## Introduction

The incidence and burden of atrial fibrillation (AF) continues to grow as the world population ages and lifespan increases. AF is a known risk factor for thromboembolic stroke and treatment with anticoagulation (AC) when medically indicated is widely accepted as standard practice^2,3^. In patients at excessive bleeding risk, occlusion of the left atrial appendage (LAA) has emerged as a means of reducing stroke risk^1,2^. The LAA is hypothesized to be a primary site of thrombus formation in AF patients due to blood stagnation within the appendage, resulting from the absence of organized left atrial contractility^1,2^. By occluding the LAA at its orifice, the potential for thrombi to emerge and enter the systemic circulation is significantly reduced^7,8^. There are several devices currently available for transcatheter LAA occlusion (LAAO) with the Watchman device and Amulet device utilized most at this time^3,5^. Following successful LAAO, post-operative imaging is required to ensure stable device positioning and absence of thrombi on the device itself^4^. Transesophageal echocardiography (TEE) is the gold standard for post-LAAO device evaluation^5^. While the timing of post-op imaging can vary, general practice is follow-up imaging at 45 days post implantation (prior to cessation of AC) and 1 year after implantation^4^. Cardiac computed tomography (CCTA) has recently emerged as a potential alternative to TEE. CCTA has several advantages from both a patient safety and a health systems standpoint^5,6,7,8^. This review aims to compare CCTA and TEE in the context of post-LAAO device implantation and to summarize the available literature comparing these two modalities.

## Methods

Articles from the databases of Medline, Pubmed, Web of Science, Cochrane, ScienceDirect, and Embase were identified. All fields in articles were searched for the following search terms. These terms were “Left Atrial Appendage Occlusion AND Transesophageal Echocardiography”, “Left Atrial Appendage Occlusion AND Cardiac Computed Tomography”, “Left Atrial Appendage Occlusion AND Transesophageal Echocardiography AND Cost”, or “Left Atrial Appendage Occlusion AND Cardiac Computed Tomography AND Cost”.

181 articles were identified in Medline. 712 articles were identified in Pubmed. 571 articles were identified in Web of Science. 232 articles were identified in Cochrane. 6054 articles were identified in Science Direct. 2435 articles were identified in Embase. The study included data from reviews and trials describing the use of TEE and CCTA for post LAAO evaluation. Data on the use of CCTA and TEE for pre- and peri-LAAO evaluation were excluded. Studies describing the use of these imaging modalities for procedures other than LAAO were excluded. Case reports, case series, editorials, and commentaries were also excluded.

## Results

Although head to head comparisons of CCTA and TEE imaging after LAAO device implant are few in number, some studies have compared the ability of these modalities to detect peridevice leaks (PDLs)^1,12,15,16,17^. One of these studies was a single center observation study of 346 patients undergoing post-operative imaging 8 weeks after LAAO with Amplatzer devices^1^. PDL was seen in 32% of patients evaluated by TEE and 61% of patients evaluated by CCTA, with this difference being statistically insignificant due to a small population size^1^. The authors also noted that leak size detected on CCTA poorly correlated with leak size seen on TEE^1^. Another study assessed the utility of CCTA in detecting complications post LAAO implantation in 45 patients^6^. Imaging was completed between 1 and 6 months after LAAO device implantation to assess for PDLs, thrombi, embolization, positioning, and pericardial effusion. PDL was noted in 64% of patients due to device malpositioning, fabric leak, and peri-device gaps, although this finding was also statistically insignifcant^6^. The most recent study comparing the two modalities utilized a cohort of 1313 patients and did identify a significant difference in PDL detection rates, with CCTA being superior to TEE^12^.

While TEE allows for detailed real-time evaluation of the LAAO device and other desired cardiac structures, there are several distinct disadvantages. First and most significant, TEE is an invasive procedure that carries some periprocedural risk including injury to the lips, teeth, tongue, or esophagus^16^. The severity of complications can range from minor soft-tissue and mucosal abrasions to esophageal perforation. Esophageal injury and perforation are the most morbid complications of TEE and also the rarest with a cited rate of 0.03-0.09% of cases^13,14^.

Procedurally, TEE is an invasive procedure that requires conscious sedation for optimal patient safety and comfort. In select higher risk cases such as patients with a high body mass index (BMI), patients with severe pulmonary hypertension, and patients with a history of active substance use disorder, sedation is provided by an anesthesiologist^16^. Once sedation is completed, the TEE probe is inserted and advanced to the midesophageal level where the study most often begins^15^. A focused assessment of the left atrium and LAA is conducted with pulse doppler signal. Once both the left atrium and LAA have been completely interrogated, the study is completed and TEE probe removed. The patient is moved to the post-procedure recovery unit while emerging from sedation^16^. Separate from probe-related mechanical injury, the conscious sedation utilized during TEE imaging also carries its own unique risks. Patients are at risk of adverse reactions to the medications used for sedation, periprocedural hemodynamic instability from the sedatives used, and potential periprocedural hypoxia and respiratory compromise^15^. While the presence of anesthesiology for periprocedural sedation helps reduce these risks, it does not completely eliminate them. Additionally, the use of a finite resource such as anesthesiology is often limited to known high risk patients, meaning that the majority of patients undergoing TEE will receive conscious sedation provided by the cardiologist and nurse performing the procedure^16^.

CCTA can play a pivotal role for pre and post procedural imaging for patients undergoing LAAO device implantation. Prior to implantation, CCTA can accurately delineate anatomical subtypes of LAA, such as the chicken wing, windsock, cactus, and cauliflower morphologies, which are important for device selection^15^. Furthermore, the LAA neck diameter and orifice shape can be precisely measured, ensuring the appropriate device is chosen. CCTA is a fast and safe procedure with a very low risk of complications. Patients with a history of iodine allergy do require premedication with steroids to ensure an allergic reaction does not occur during the study although rates of such complications are uncommon and not readily found^15^. Moreover, intravenous iodinated contrast has been linked to transient worsening of renal function and acute kidney injury in certain patients. Thus in the cohort of patients with a history of chronic kidney disease, the use of CCTA may be limited or restricted based on the calculated glomerular filtration rate at time of study^16^. Finally, CCTA utilizes x-ray radiation for image acquisition which has been linked to an elevated risk of cancer over time^16^. The amount of radiation delivered during a CCTA is variable and based on the protocol used at each respective institution. Literature cites radiation rates of 3-14mSv for a CCTA; for comparison, a chest x-ray has an approximate radiation dose of 0.03mSv^17^.

In a financial perspective, the costs of using CCTA and TEE have been alluded to in a few studies. Although data pertaining to the expenses of these imaging modalities are highly institution specific, certain studies have done cost analyses and demonstrated that expenditures range from $14,000 to $32,000 depending on the equipment and personnel utilized^18,19^. However, these analyses were performed in the context of peri-procedural LAAO evaluation and non LAAO evaluations. Thus the true expenses associated with each modality in post LAAO evaluation are an area for further investigation.

## Discussion

As TEE is the current gold standard for post LAAO imaging, cardiologists are able to assess for device positioning and thrombi with a combination of two and three dimensional images^1^. Additionally, doppler signals can be utilized to assess for peridevice flow and leakage, indicating a poor seal of the appendage^1,5^. Given the realtime nature of TEE, cardiologists are able to augment probe positioning and image acquisition characteristics in realtime to generate the highest quality and most clinically informative images possible^3^. Additionally, cardiologists can assess other components of cardiac function such as biventricular function and valve function during the study^3^.

From a systematic standpoint, TEE is a fairly resource intensive procedure. A cardiologist, procedural nurse, procedure suite, and staffed post-procedure care unit are required to perform a TEE^15,17^. In complex patients, an anesthesiologist is also often present to aid with administration of intra-procedural sedation and hemodynamic monitoring. While the procedure itself often takes less than one hour to complete, the setup and post-procedure monitoring and care all add time and cost to the overall study time^16^.

CCTA meanwhile has emerged as an alternative imaging modality in the evaluation and management of LAAO devices following implantation. CCTA utilizes X-ray technology with iodinated contrast material to produce high-resolution images of the heart and its associated vasculature^3,6^. The use of multi-detector row CT scanners has significantly improved image quality and temporal resolution, enabling accurate evaluation of cardiac structures and devices. Advanced CT post-processing techniques enable the creation of virtual device simulations, providing valuable information for predicting device behavior and implantation success^11,12^. Additionally CCTA is instrumental in evaluating the optimal access sites, such as the transseptal or transthoracic routes, which are critical for successful device deployment^16^. Following device implantation, CCTA can be utilized as a viable alternative to TEE for post-procedure assessment of LAAO devices. CCTA allows for accurate detection of peri-device leaks, which can occur as a result of incomplete sealing and may lead to thromboembolic complications^16^.

Furthermore, CCTA enables the identification of device thrombus formation, which can also increase the risk of thromboembolic events^11,12^. Timely detection of these complications is crucial for the prompt initiation of appropriate management strategies. Additionally, CCTA provides valuable information on device positioning and stability within the LAA, ensuring that the device is functioning effectively and securely anchored to the appendage^17^. This information is essential for monitoring device performance and guiding further management if required.

## Limitations

Although CCTA has shown superiority over TEE in detection of PDLs, PDL size, and LAATs, few of these findings have been established as significantly different between the modalities. This can be attributed to the small size of the population in which post LAAO imaging has been performed, thus making it more challenging to establish power and a true difference between the two imaging modalities. Data pertaining to the financial aspects of using these imaging modalities were also limited in the literature. Ultimately, the clinical and economic aspects of CCTA vs TEE warrant further investigation for a determination as to which modality would be suitable for post LAAO implant evaluation.

## Conclusions

CCTA has some key advantages over TEE in post LAAO evaluation such as higher rates of detection for PDLs, LAATs, and device positioning. CCTA has ease and speed of performance, requiring less manpower for completion, although at the price of radiation exposure. Future investigations will help determine if CCTA has a significantly higher rate of complication detection compared to TEE. Additional investigations will also determine the patient level outcomes and cost benefit of each modality.

## Ethical Statement

Ethical/IRB approval for our submission was not obtained as patient data was not involved.

## Data Availability

All data produced in the present work are contained in the manuscript

## Abbreviations

LAA: left atrial appendage,
LAAO: left atrial appendage occlusion,
TEE: transesophageal echocardiography,
CCTA: cardiac computerized tomography angiography,
PDL: peri-device leak,
CKD: chronic kidney disease,
LAAT: left atrial appendage thrombus,
AF: atrial fibrillation

